# Predicting mortality due to SARS-CoV-2: A mechanistic score relating obesity and diabetes to COVID-19 outcomes in Mexico

**DOI:** 10.1101/2020.04.20.20072223

**Authors:** Omar Yaxmehen Bello-Chavolla, Jessica Paola Bahena-López, Neftali Eduardo Antonio-Villa, Arsenio Vargas-Vázquez, Armando González-Díaz, Alejandro Márquez-Salinas, Carlos A. Fermín-Martínez, J. Jesús Naveja, Carlos A. Aguilar-Salinas

## Abstract

**BACKGROUND:** The SARS-CoV-2 outbreak poses challenge to healthcare systems due to high complication rates in patients with cardiometabolic diseases. Here, we identify risk factors and propose a clinical score to predict COVID-19 lethality, including specific factors for diabetes and obesity and its role in improving risk prediction.

**METHODS:** We obtained data of confirmed and negative COVID-19 cases and their demographic and health characteristics from the General Directorate of Epidemiology of Mexican Ministry of Health. We investigated specific risk factors associated to COVID-19 positivity and mortality and explored the impact of diabetes and obesity on modifying COVID-19 related lethality. Finally, we built a clinical score to predict COVID-19 lethality.

**RESULTS:** Among 177,133 subjects at May 18^th^, 2020, we observed 51,633 subjects with SARS-CoV-2 and 5,332 deaths. Risk factors for lethality in COVID-19 include early-onset diabetes, obesity, COPD, advanced age, hypertension, immunosuppression, and CKD; we observed that obesity mediates 49.5% of the effect of diabetes on COVID-19 lethality. Early-onset diabetes conferred an increased risk of hospitalization and obesity conferred an increased risk for ICU admission and intubation. Our predictive score for COVID-19 lethality included age ≥65 years, diabetes, early-onset diabetes, obesity, age <40 years, CKD, hypertension, and immunosuppression and significantly discriminates lethal from non-lethal COVID-19 cases (c-statistic=0.823).

**RESULTS:** Here, we propose a mechanistic approach to evaluate risk for complications and lethality attributable to COVID-19 considering the effect of obesity and diabetes in Mexico. Our score offers a clinical tool for quick determination of high-risk susceptibility patients in a first contact scenario.

## INTRODUCTION

The first cases of SARS-CoV-2 infection in Mexico were reported at the end of February [1]; since then, the number of COVID-19 cases has been steadily increasing, with most fatal cases being associated with the presence of comorbidity and, particularly, cardiometabolic comorbidities. A high prevalence of cardiometabolic diseases worldwide represents a challenge during the COVID-19 epidemic; an elevated number of patients with SARS-CoV-2 infection have a preexisting disease such as obesity, hypertension, cardiovascular disease, diabetes, chronic respiratory disease or cancer [2,3]. Diabetes mellitus and obesity represent a large share of the cardiometabolic morbidity burden of the region [4]; moreover, most cases of diabetes remain either undiagnosed or lack adequate glycemic control, posing them at risk of increased COVID-19 severity. Despite several reports evaluating the burden of comorbidities including obesity, diabetes and hypertension on the clinical course of COVID-19, the joint role of obesity and diabetes in modifying COVID-19 outcomes has not been fully explored [5].

Several studies have demonstrated a higher susceptibility to acute respiratory infectious diseases in people with diabetes [6]. Moreover, diabetes and obesity have been described as independent risk factors for severe pulmonary infection [7,8]. Obesity influences the clinical outcomes during acute severe respiratory distress syndrome (ASRDS); obesity has been proposed as a protective factor for mortality following lung injury due to reverse causality or as a cause of mortality and adverse clinical outcomes for severe influenza cases due to mechanical and immunologic factors. In the cases of the COVID-19 outbreak, obesity has been consistently associated with adverse outcomes [9,10]. Furthermore, a large proportion of obesity cases in Mexico live in geographical areas of increased social vulnerability, which poses a structural inequality that might also increase mortality for COVID-19 associated to both diabetes and obesity [11]. Chronic inflammation in obesity might worsen the acute inflammatory response triggered by SARS-CoV-2 infection, which might be associated to a cytokine release syndrome [5,12]. Here, we investigate the role of both diabetes and obesity in determining propensity for SARS-CoV-2 infection and its associated clinical outcomes including disease severity and COVID-19 lethality; using these associations, we further construct a clinically useful predictive model for COVID-19 mortality using national epidemiological surveillance data from Mexico.

## METHODS

### Data sources

We extracted data from the General Directorate of Epidemiology of the Mexican Ministry of Heath, which is an open-source dataset comprising daily updated information of suspected COVID-19 cases which have been confirmed with a positive test for SARS-CoV-2 certified by the National Institute for Diagnosis and Epidemiological Referral [13].

### Definitions of suspected and confirmed COVID-19 cases

The Ministry of Health defines a suspected COVID-19 case as an individual of any age whom in the last 7 days has presented cough, fever or headache (at least two), accompanied by either dyspnea, arthralgias, myalgias, sore throat, rhinorrhea, conjunctivitis or chest pain. Amongst these suspected cases, the Ministry of Health establishes two protocols for case confirmation: 1) SARS-CoV-2 testing is done widespread for suspected COVID-19 cases with severe acute respiratory infection with signs of breathing difficulty or deaths in suspected COVID-19 cases, 2) for all other suspected cases, a sentinel surveillance model is being utilized, whereby 475 nationally representative health facilities sample ~10% of mild outpatient cases and all suspected severe acute respiratory infection [14]. Demographic and health data are collected and uploaded to the epidemiologic surveillance database by personnel from the corresponding individual facility.

### Variables and definitions

Available information for all confirmed, negative and suspected COVID-19 cases includes age, sex, nationality, state and municipality where the case was detected, immigration status as well as identification of individuals who speak indigenous languages. Health information includes status of diabetes, obesity, chronic obstructive pulmonary disease (COPD), immunosuppression, pregnancy, arterial hypertension, cardiovascular disease, chronic kidney disease (CKD) and asthma. Date of symptom onset, hospital admission and death are available for all cases as well as treatment status (outpatient or hospitalized), information regarding diagnosis of pneumonia, ICU admission and whether the patient required invasive mechanical ventilation. Early-onset diabetes was defined as a medically diagnosed case of diabetes mellitus in subjects younger than age 40 years. The majority of early-onset diabetes cases are patients with type 2 diabetes. This phenotype is common in Mexico; it is characterized for having a more aggressive form of the disease usually associated with obesity, rapidly declining β-cell function, higher risk of microvascular complications compared to late-onset type 2 diabetes [15]. We considered this form of diabetes given its high prevalence in Mexican and other populations as well as its higher propensity for complications [16,17].

### Statistical analysis

#### Comorbidities associated to SARS-CoV-2 positivity

We investigated the association of demographic and health data associated with SARS-CoV-2 positivity using logistic regression analyses, excluding individuals who were only suspected but unconfirmed cases of COVID-19. Next, we stratified these analyses for individuals with only diabetes or only obesity to identify specific risk factors within these populations, especially focusing on individuals who were <40 years and likely acquired the disease early.

#### COVID-19 mortality risk

In order to investigate risk factors predictive of COVID-19 related 30-day lethality, we fitted Cox Proportional risk regression models estimating time from symptom onset up to death or censoring, whichever occurred first in cases with confirmed positivity for SARS-CoV-2. To identify diabetes and obesity-specific risk factors, we carried out stratified analyses. Given the availability of SARS-CoV2 negative cases within the dataset, we fitted Cox models for mortality which included SARS-CoV-2 positivity as an interaction term with different comorbidities, hypothesizing that some factors increase mortality risk specifically for COVID-19. Finally, we fitted a logistic regression model only for mortality cases to evaluate associations with lethality rates in COVID-19 related and non-related deaths.

#### Influence of obesity and diabetes in COVID-19 related outcomes

Finally, we estimated factors associated to admission to hospital facilities, intensive care units (ICUs) and requirements for mechanical ventilation in all confirmed COVID-19 cases using logistic regression. To identify specific factors for all COVID-19 and non-COVID-19 patients related to outcomes, we included interaction effects with comorbidities factors; we also performed Kaplan-Meier analyses to identify the role of comorbidities in modifying lethality risk in individuals with diabetes and obesity and compared across categories using Breslow-Cox tests. Finally, we performed causal-mediation analyses with causally-ordered mediators using a previously validated approach to investigate whether obesity mediates the decreases in COVID-19 survival attributable to diabetes, particularly in early-onset cases <40 years [18].

#### Mechanistic mortality risk score for COVID-19

Finally, we constructed a clinically useful model to predict lethality in COVID-19 cases which might be useful to apply in first contact settings, including variables and interactions which were identified in mortality analyses. The model was trained in 80% of the dataset split using random sampling stratified by mortality status with the *caret* R package and was later validated in the remaining 20%. Points were assigned by standardizing all β coefficients with the minimum absolute β coefficient obtained from Cox regression. Points were stratified according to categories of Low risk (≤0), Mild risk (1-3), Moderate risk (4-6), High risk (7-9) and very high risk (≥10). Risk across categories was verified using Kaplan-Meier analyses. C-statistics and D_xy_ values were corrected for over-optimism using k-fold cross-validation (k=10) using the *rms* R package. A p-value <0.05 was considered as statistical significance threshold. All analyses were performed using R software version 3.6.2.

## RESULTS

### COVID-19 cases in Mexico

At the time of writing this report (May 18^th^, 2020), a total of 177,133 subjects had been treated initially as suspected COVID-19 cases. Amongst them, 51,633 had been confirmed as positive and 98,567 tested negative for SARS-CoV-2 infection; additionally, 26,933 cases were still being studied as suspected cases pending testing reports. Amongst confirmed cases, 5,332 deaths were reported (10.33%) whilst 2,009 deaths were SARS-CoV-2 negative cases (2.04%) and 656 deaths of suspected but unconfirmed cases (2.44%) had been reported. Compared to SARS-CoV-2 negative cases, confirmed cases were older, predominantly male (1.37:1 ratio), had higher rates of hospitalization and showed a higher prevalence of diabetes, hypertension and obesity. SARS-CoV-2 cases were also more likely have higher rates of ICU admission and requirements for invasive ventilation compared to negative cases (**Table 1**).

**Table 1.** Descriptive statistics comparing negative, positive and suspected cases for SARS-CoV-2 in Mexico at 04/27/2020. *Abbreviations*: ICU= intense care unit; COPD= chronic obstuctive pulmonary disease; CKD= chronic kidney disease; CVD= cardiovascular disease.

| Parameter | Positive for SARS-CoV-2<br>n=51633 | Negative for SARS-CoV-2<br>n=98567 | Suspected for SARS-CoV-2<br>n=26933 |
| --- | --- | --- | --- |
| Age (mean ± sd) | 46.65 ± 15.83 | 40.25 ± 17.33 | 43.3 ± 16.55 |
| Male sex (%) | 29803(57.7) | 47177(47.9) | 13602(50.5) |
| Mortality (%) | 5332(10.3) | 2009(2.0) | 656(2.4) |
| Hospitalization (%) | 19831(38.4) | 18586(18.9) | 6674(24.8) |
| Pneumonia (%) | 14919(28.9) | 11928(12.1) | 4723(17.5) |
| ICU admission (%) | 1893(3.7) | 1456(1.5) | 464(1.7) |
| Invasive ventilation (%) | 1959(3.8) | 1162(1.2) | 406(1.5) |
| Diabetes (%) | 9460(18.3) | 10553(10.7) | 3442(12.8) |
| COPD (%) | 1131(2.2) | 2238(2.3) | 444(1.6) |
| Asthma (%) | 1602(3.1) | 4530(4.6) | 725(2.7) |
| Immunosuppression (%) | 849(1.6) | 2472(2.5) | 393(1.5) |
| Hypertension (%) | 11151(21.6) | 14858(15.1) | 4453(16.5) |

| Parameter | Positive for SARS-CoV-2 | Negative for SARS-CoV-2 | Suspected for SARS-CoV-2 |
| --- | --- | --- | --- |
|  | n=51633 | n=98567 | n=26933 |
| Obesity (%) | 10708(20.7) | 14011(14.2) | 4364(16.2) |
| CKD (%) | 1265(2.4) | 2270(2.3) | 525(1.9) |
| CVD (%) | 1381(2.7) | 2891(2.9) | 595(2.2) |
| Smoking (%) | 4366(8.5) | 9624(9.8) | 2451(9.1) |

### Factors associated with COVID-19 positivity

We investigated cases related to COVID-19 positivity within Mexico. We found that odds of SARS-CoV-2 positivity was higher with diabetes, hypertension, obesity, age >65 and male sex. When assessing age, we observed reduced odds of SARS-CoV-2 positivity in patients <40 years and, in contrast, when exploring its interaction with diabetes, we observed an increased probability of SARS-CoV-2 infection. In stratified models, we observed that for patients with diabetes, SARS-CoV-2 positivity was associated with obesity and male sex; for patients with obesity, diabetes and male sex were also significant as was the interaction between diabetes and age <40 years [19].

### Predictors for COVID-19 related 30-day mortality

We identified that COVID-19 cases were associated with a near four-fold increase in mortality due to acute respiratory infection (HR 3.967, 95%CI 3.739-4.210) compared to non-COVID-19 cases. Of interest, the only comorbidity which conferred increased lethality risk exclusively for COVID-19 compared to non-COVID-19 was obesity (HR 1.261, 95%CI 1.109-1.433, **Figure 1A**). Factors associated to increased lethality in COVID-19 cases were age >65 years, diabetes mellitus, obesity, CKD, COPD, immunosuppression and hypertension, whilst asthma showed a protective effect (**Figure 1B**). We searched for an interaction between diabetes mellitus and age <40 years to account for early-onset diabetes mellitus, adjusted for sex and obesity; a higher mortality risk was found for early-onset diabetes cases (HR 2.754, 95%CI 2.259-3.359). Adjusting the model for pneumonia to account for SARS-CoV-2 severity as a predictor of mortality (HR 5.264, 95%CI 4.933-5.618), we observed that asthma was no longer associated to decreased mortality, whilst all other predictors remained constant.

**Figure 1.**
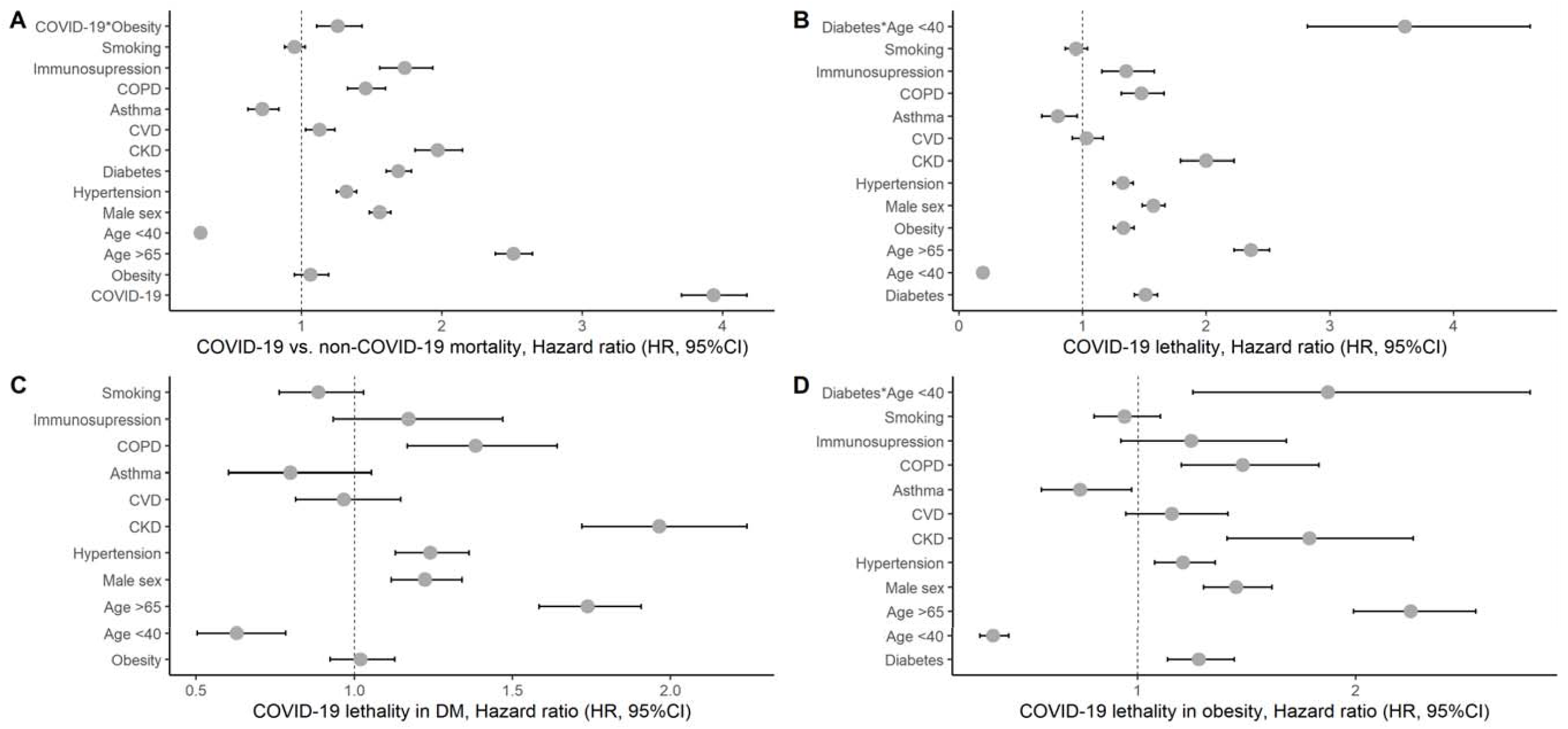
Cox proportional risk regression analysis to evaluate lethality of SARS-CoV-2 in Mexico, compared to SARS-CoV2 negative cases for all suspected cases with SARS-CoV2 status available (A) and stratified by diabetes mellitus (B) and obesity (C). *Abbreviations*: ICU= intense care unit; COPD= chronic obstructive pulmonary disease; CKD= chronic kidney disease; CVD= cardiovascular disease, HR= Hazard ratio.

### COVID-19 in patients with diabetes mellitus

Confirmed COVID-19 cases with diabetes had a mean age of 57.16 (±12.83) years and were predominantly male. This population had particularly higher mortality rate (21.8% vs. 7.7%), hospitalization, ICU admission, requirement for invasive ventilation and confirmed pneumonia compared with those without diabetes. When stratifying mortality, those with early onset diabetes (<40 years) had higher mortality rates compared to individuals <40 years without diabetes (11.3% vs. 1.3%); similarly, those aged >40 years without diabetes had lower rates compared to those >40 years with diabetes (12.0% vs. 22.7%). As expected, obesity, hypertension, COPD, CKD, CVD, and immunosuppression were also more prevalent in this population. [19]. In patients with diabetes mellitus, COVID-19 related mortality was higher in those with concomitant immunosuppression, COPD, CKD, hypertension and those aged >65 years (**Figure 1C)**. When assessing the role of comorbidities, COVID-19 patients with diabetes, coexistent obesity, those with early onset diabetes (<40 years) or an increase in the number of comorbidities had increased risk of COVID-19 lethality (Breslow test p<0.001, **Figure 2**). When comparing cases with and without COVID-19 amongst patients with diabetes mellitus we observed a threefold higher risk of mortality associated with COVID-19 (HR 3.375, 95%CI 3.103-3.672) after adjustment for age, sex and comorbidities [19].

**Figure 2.**
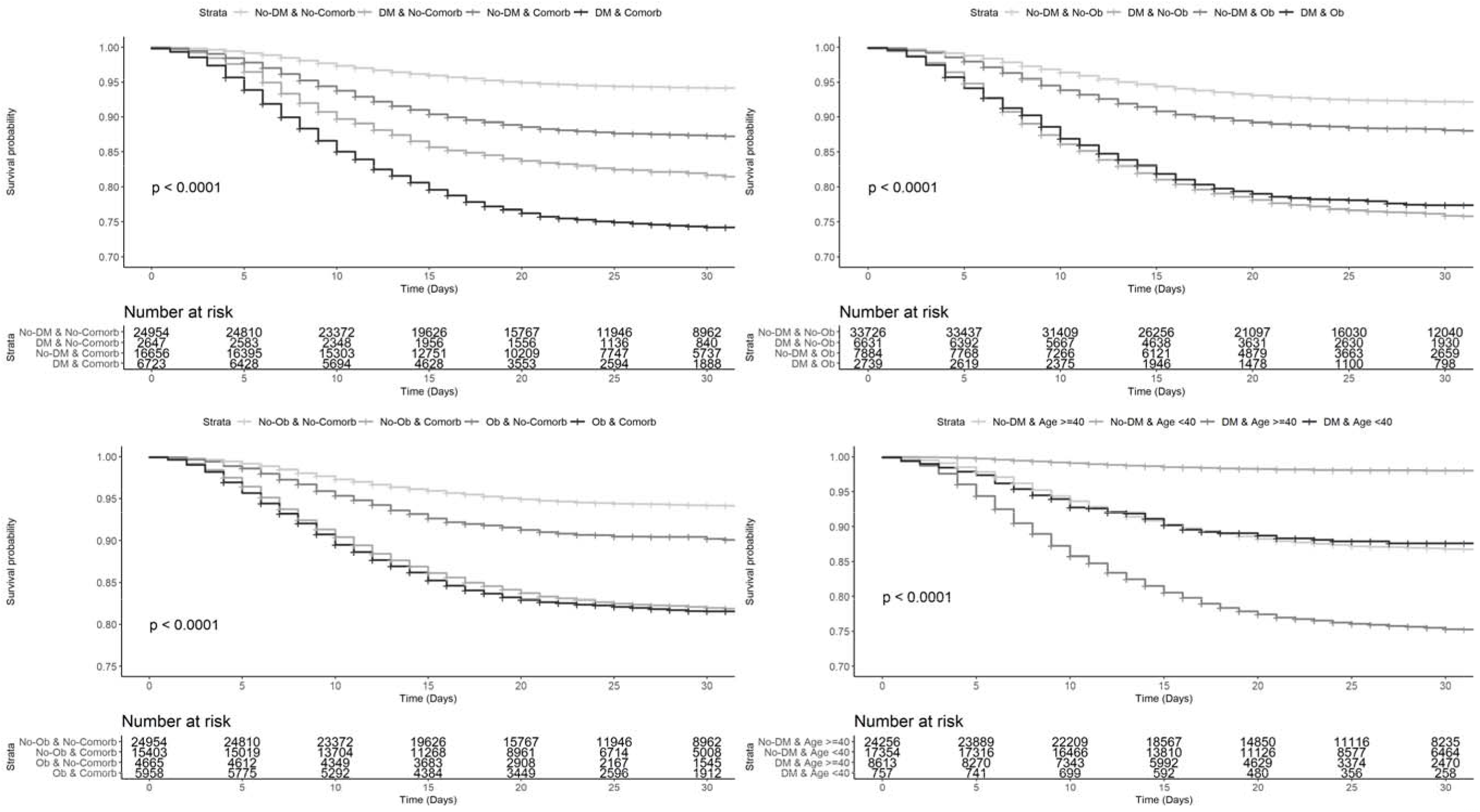
Kaplan-Meier survival curves to evaluate lethality of SARS-CoV-2 positivity in patients with diabetes and comorbidities (A), diabetes and obesity (B), obesity and comorbidities (C) and diabetes with age 40 years (D). Abbreviations: DM= Diabetes mellitus; OB= Obesity; Comorb= Comorbidities.

### COVID-19 in patients with obesity

Similar to patients with diabetes, confirmed COVID-19 cases with obesity had particularly higher rates of mortality (13.5% vs. 9.4%), hospitalization and confirmed pneumonia. Furthermore, patients with obesity also had higher rates of ICU admission (5.0% vs. 3.3%) and were more likely to be intubated (5.2% vs. 3.3%). As expected, diabetes, hypertension, COPD, smoking, CVD and asthma were also more prevalent in patients with obesity [19]. As previously mentioned, obesity was identified as a risk factor which displayed differential risk for COVID-19 infection, specific risk factors for lethality in obese patients with COVID-19 infection included COPD, CKD, hypertension, male sex, age >65, diabetes and, particularly, early-onset type 2 diabetes (**Figure 1D**); overall, COVID-19 increased risk of mortality in obesity nearly five-fold (HR 4.989, 95%CI 4.444-5.600). The addition of obesity to any number of comorbidities significantly increased the risk for COVID-19 lethality (Breslow test p<0.001, **Figure 2-C**). Using causally ordered mediation analysis we investigated whether the effect of diabetes (E) on COVID-19 related lethality (Y) was partially mediated by obesity (M); the direct effect of diabetes on COVID-19 lethality was significant (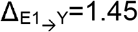, 95%CI 1.36-1.54) as was the indirect effect of obesity (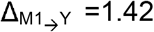, 95%CI: 1.36-1.49), representing 49.5% of the total effect of diabetes.

### COVID-19 outcomes and comorbidities

Given the increased risk of diabetes and obesity in modifying COVID-19 related lethality, a secondary objective of our study was to investigate its associations with inpatient outcomes, including hospitalization rate, ICU admission and requirement for mechanical ventilation. In general, early-onset diabetes patients and those with obesity had higher risk of hospitalization, whilst patients with obesity also had increased risk for ICU admission and required intubation. Patients with diabetes mellitus overall had higher risk of hospitalization, risk of ICU admission and intubation (**Figure 3**).

**Figure 3.**
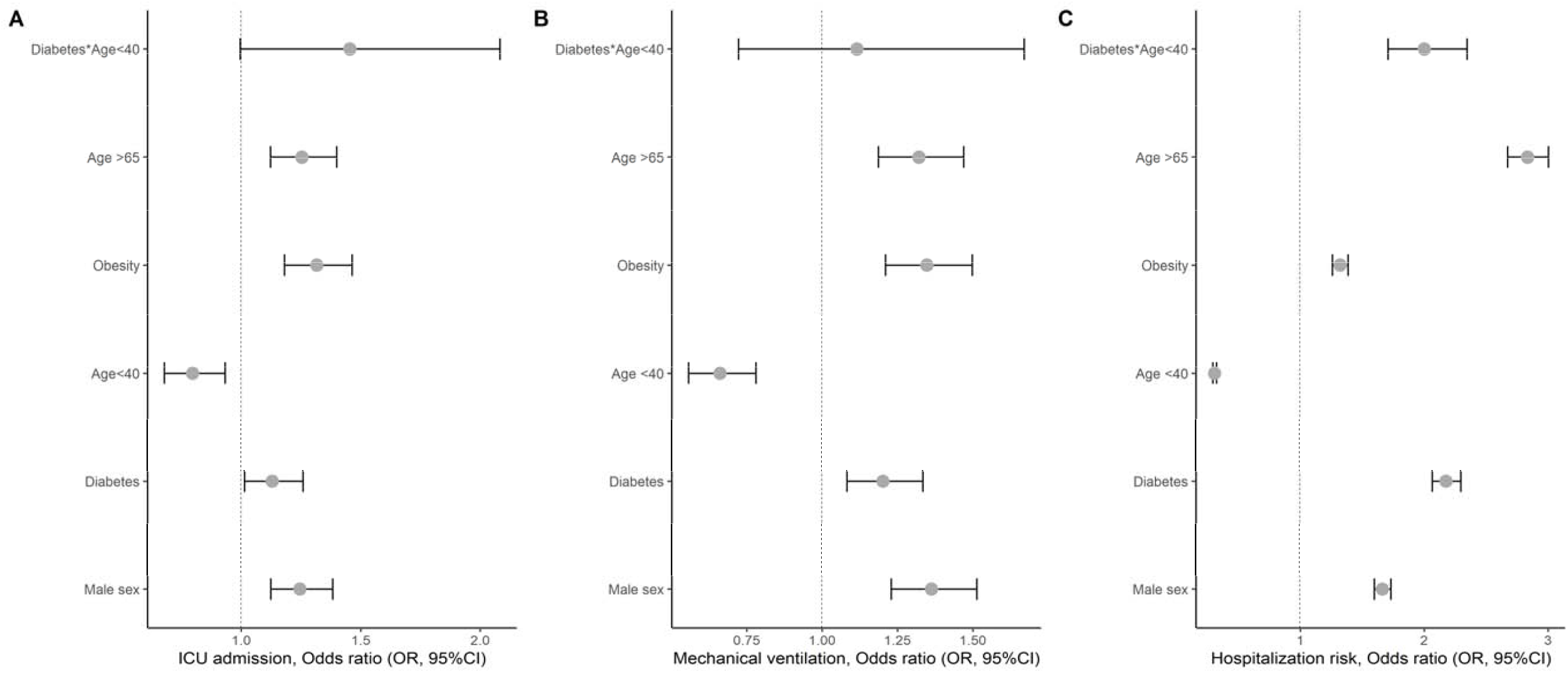
Logistic regression analyses to evaluate COVID-19 related outcomes in all patients with SARS-CoV2 positivity for admission to ICU (A), mechanical ventilation (B) and hospital admission risk (C).

### Mechanistic score for mortality in COVID-19

Using the identified predictors for mortality and the observed interaction for early-onset diabetes, we designed a predictive score for COVID-19 mortality using Cox regression with a random split of 80% of the dataset stratified by mortality (n=41,307, deaths=4,276). We identified as significant predictors age>65 years, diabetes mellitus, obesity, CKD, COVID-19 related pneumonia, COPD, and immunosuppression (**Table 2**); age<40 was a protective factor which was modified by its interaction with T2D (R^2^=0.154, c-statistic= 0.817, D_xy_=0.647); assigning the point system did not significantly reduce the model’s performance (R^2^=0.154, C-statistic=0.822, D_xy_=0.645). Finally, category stratification reduced only moderately performance statistics (R^2^=0.152, C-statistic=0.810, D_xy_=0.620), and were not significantly modified after cross-validation correction (R^2^=0.167, D_xy_=0.645). The score was then validated using the remaining 20% of the population (n=10,326, deaths=1,056); we observed that the score retained its predictive and discriminative ability (R^2^=0.167, C-statistic=0.830, D_xy_=0.660) as did the categories (R^2^=0.170, C-statistic=0.821, D_xy_=0.642). Distribution of the score significantly discriminates between lethal and non-lethal COVID-19 cases (**Figure 4**).

**Table 2.**
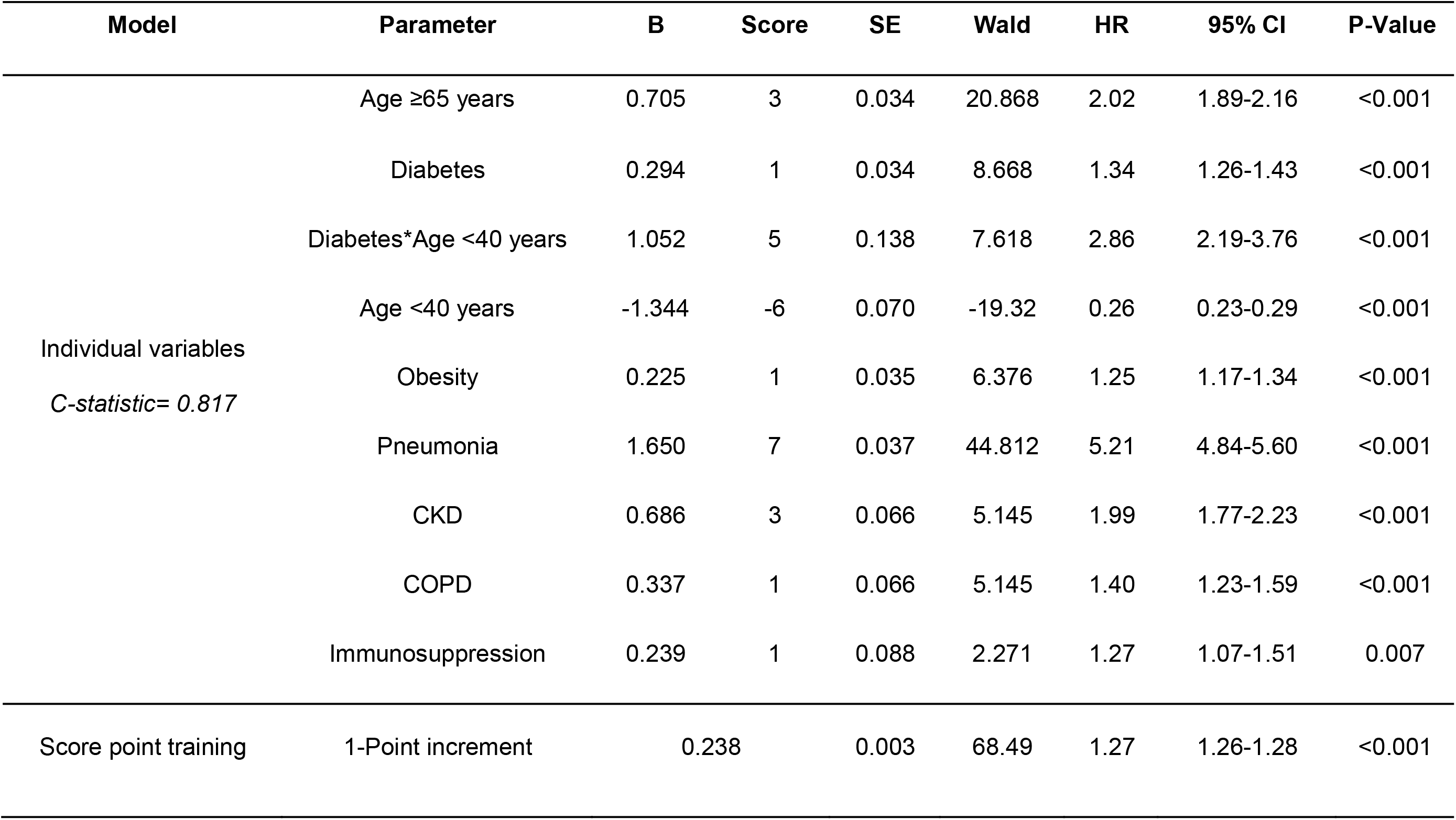
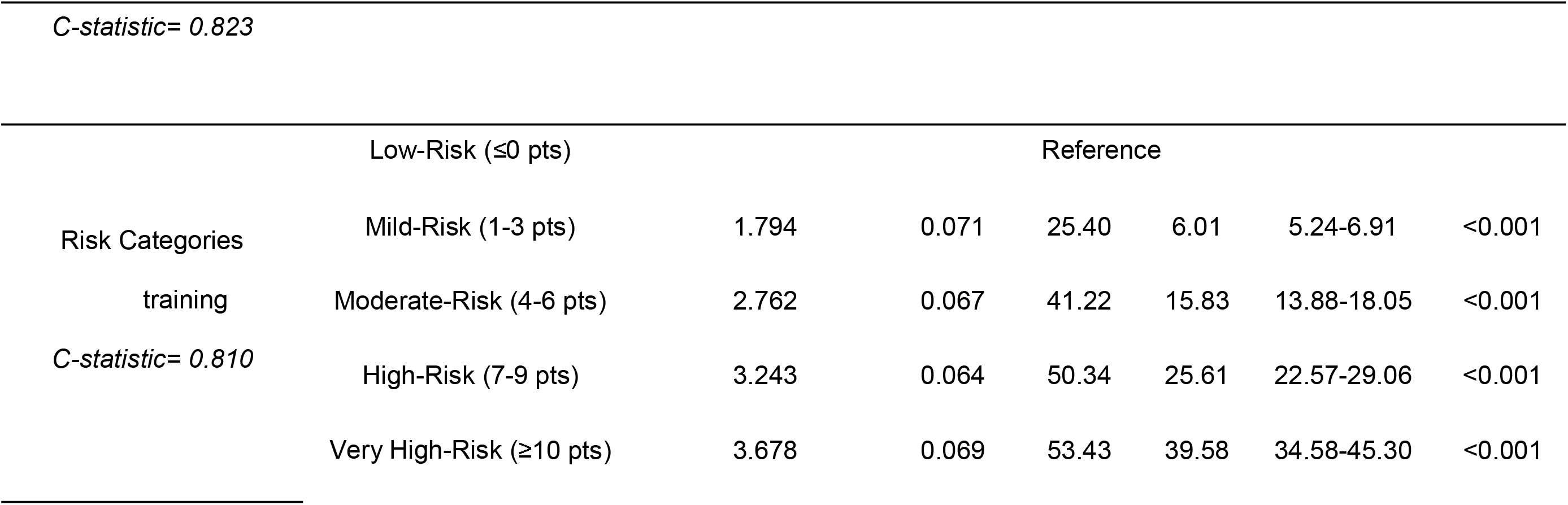
Cox proportional risk models for lethality using the mechanistic COVID-19 lethality score in confirmed cases of COVID-19 using individual components, single score point and risk stratification categories. Abbreviations: COPD= chronic obstructive pulmonary disease; CKD= chronic kidney disease; CVD= cardiovascular disease.

**Figure 4.**
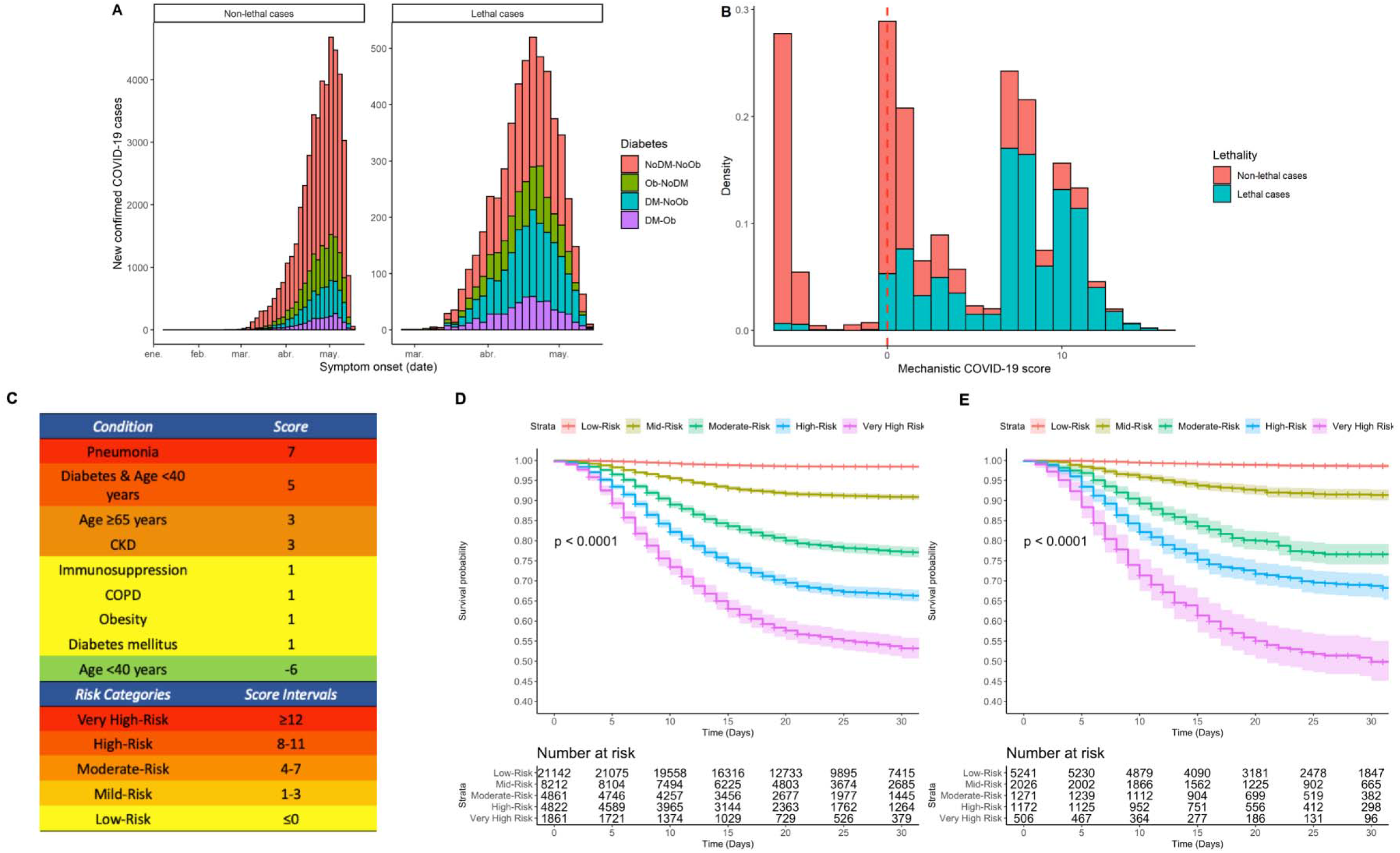
Symptom onset among lethal and non-lethal cases in new-confirmed COVID 19 cases, stratified by diabetes and obesity status (A), density histogram of scores of the mechanistic COVID-19 score (B). Points and score intervals considered for clinical score scale, where Diabetes & Age<40 represent the score of the interaction term (C) and Kaplan-Meir Survival analysis curves to evaluate lethality using risk categories in the training (D) and validation cohorts (E). Abbreviations: OB= Obesity; DM= Diabetes mellitus; CKD= chronic kidney disease; COPD= Chronic obstructive pulmonary disease.

## DISCUSSION

Our results demonstrate that diabetes, particularly early-onset diabetes, obesity and comorbidity burden modify risk profiles in patients with COVID-19 in Mexico and significantly improve mortality prediction related to COVID-19 lethality. These findings position the notion that early-onset type 2 diabetes might carry a higher risk of mortality in younger patients and the risk is similar to older patients with other comorbidities and only higher in older patients with diabetes. Inclusion of the interaction term (Diabetes*Age <40 years) within the risk score effectively offsets the protective effect of younger age, indicating the higher risk attributable to early-onset diabetes and the utility of including this term within the risk score to improve prediction of mortality. Furthermore, our results suggest that obesity is a COVID-19 specific risk factor for mortality, risk of ICU admission, tracheal intubation and hospitalization and even increases risk in patients with comorbid diabetes and COVID-19 infection. Overall, this positions the co-existence of obesity and diabetes, particularly early-onset diabetes, as a considerable risk factor for COVID-19 mortality in Mexicans, whom have reported an alarmingly high burden of both conditions in recent health surveys.

The relationship between increased risk of mortality attributable to acute severe respiratory infections in patients with diabetes mellitus has been extensively reported, particularly for the acute respiratory syndrome caused by SARS-CoV-1 [20-22]. Evidence relating SARS-CoV-2 infections in China demonstrated increased rates of diabetes mellitus in hospitalized patients and in those with increased disease severity as assessed by ICU admission and requirement for invasive ventilation. Additionally, hospitalized patients have shown increased rates of both obesity and diabetes for COVID-19 compared to non-hospitalized cases in the US, China and Italy [5,23,24]. Increased susceptibility for COVID-19 in patients with diabetes may be explained for several potential mechanisms including an increased lung ACE2 expression and elevated circulating levels of furin, a protease involved in viral entry to cells, and a decreased clearance of SARS-CoV-2 viral particles in subjects with diabetes and/or hypertension associated with ACE2 expression [25-28]. Impairments in immunity observed in patients with diabetes are characterized by initial delay in activation of Th1 cell-mediated immunity and late hyper-inflammatory response and are consistent with the increased risk associated with additional immunosuppression observed with our data [29]. Additional factors which have been proposed to modify COVID-19 mortality risk and worsen glycemic control in diabetes include corticosteroid therapy, inadequate glucose monitoring, the effect of social distancing on diabetes care and the use of antihypertensive medication; however these factors remain to be confirmed by clinical evidence [30]. Given the large proportion of undiagnosed diabetes cases in Mexican and poor glycemic control reported by recent estimates, the burden of COVID-19 might be higher than expected in Mexico and poses a challenge for the Mexican healthcare system to give particular attention to this sector as the epidemic moves forward [31-33].

Diabetes mellitus is one of the main causes of morbidity and it accounts for a large proportion of mortality risk in Mexican population [4]. Of relevance, Mexicans have increased risk of diabetes and diabetes-related obesity attributable to genetic variants associated to its Amerindian ancestry, and an earlier age of onset independent of body-mass index [34,35]. Data on the incidence of early-onset type 2 diabetes in Mexican population position obesity and insulin resistance as significant risk factors, which are also highly prevalent in younger patients and increase metabolic risk [16,36]. These associations partly explain the increased risk of COVID-19 lethality in younger patients within our cohort despite the younger average age of Mexican population and poses early-onset diabetes mellitus as a significant risk factor for COVID-19 mortality and increased severity of infection in younger patients [5,37].

In our work, we demonstrate that compared to non-COVID-19 infections, obesity significantly modifies the risk of mortality attributable to COVID-19 infection. Obesity and, in particular, abdominal obesity, is one of Mexico’s main public health problems; in recent years the socio-economic burden of obesity as well as its impact on mortality have increased drastically, with the Ministry of Health declaring a state of epidemiological emergency [38]. Evidence from different regions has supported the notion that obesity increases mortality risk and severity of COVID-19 infections, which holds particularly true for younger patients [2,39]. Obesity is characterized by low-grade inflammation, whereby mononuclear cells increase transcription of pro-inflammatory cytokines; obesity also interacts with insulin resistant states and metabolic syndrome traits often comorbid in subjects with obesity to further promote inflammatory and a pro-thrombotic states which might lead to deleterious responses to infectious pathogens [40,41]. Furthermore, obesity has been shown to lead to decrease immune response to infectious pathogens which in turn also may affect the lung parenchyma, increasing the risk for inflammatory lung diseases of infectious causes, like influenza and SARS-CoV-2 [42-44]. Similar inflammatory responses have been attributed to the low number of asthmatics with confirmed SARS-CoV-2 infection. Immune Th2 response observed in asthma may counter inflammation related to SARS-CoV-2 infection, as was previously reported for cases in Wuhan; however, increased pro-inflammatory processes in severe forms of COVID-19 likely outweigh the effect of asthma. Additionally, abdominal obesity reduces the compliance of lung, chest wall and the entire respiratory system, resulting in impaired ventilation of the base of the lungs and reduced oxygen saturation of blood [45]. Recently, Simonnet et al. explored the high prevalence of obesity in patients with COVID-19 reporting that obesity is a risk factor for SARS-CoV-2 infection severity independent of age, diabetes and hypertension. Notably, ACE2 expression in adipose tissue is higher than in the lung and its expression profile is not different in obese and non-obese subjects; however, obese subjects have more adipocytes; thus, they have a greater number of ACE2-expressing cells and thus higher likelihood of SARS-CoV-2 entry [2,46]. Our data shows that obesity is a specific risk factor for COVID-19 related outcomes and that it partly mediates the risk associated with diabetes mellitus. Public health efforts by the Mexican government in epidemiological surveillance have largely focused in identifying patient’s at highest risk of complications; these findings could inform public health decisions and increase awareness on the role of obesity in modifying risk of COVID-19 outcomes.

Our study had some strengths and limitations. First, we analyzed a large dataset which included information on both confirmed positive and negative SARS-CoV-2 cases, which provides a unique opportunity to investigate COVID-19 specific risk factors and develop a predictive model for COVID-19 mortality. Additionally, with the database being nationally-representative it allows for reasonable estimates on the impact of both diabetes and obesity despite the possibility of important regional differences in cardiometabolic risk which might influence risk estimates. A potential limitation of this study is the use of data collected from a sentinel surveillance system model, which is skewed towards investigating high risk cases or only those with specific risk factors which on one hand increases power to detect the effect of comorbidities and on the other hand might not be representative of milder cases of the disease; this is demonstrated in the risk of COVID-19 positivity, which is higher for high risk cases. The updating daily estimates of COVID-19 cases are unlikely to change the direction of the identified associations though it might modify numeric estimates. The role of a risk-gradient related to BMI and increasing degrees of obesity could not be explored with available data and remains as an area to be explored in further studies. Implementation of our proposed model might be useful to allocate prompt responses to high risk cases and improve stratification of disease severity.

In conclusion, we show that both diabetes and obesity increase the risk of SARS-CoV-2 infection in Mexico. In particular, diabetes increases the risk of COVID-19 related mortality and, specifically, increases mortality risk in early-onset cases. Obesity is a COVID-19 specific risk factor for mortality and for increased disease severity; obesity also is a partial mediator on the effect of diabetes in decreasing survival associated with COVID-19 infection. This mechanistic interpretation on the risk of comorbidities allowed the development of a model with good performance to predict mortality in COVID-19 cases. Given the burden of obesity and diabetes in Mexico, COVID-19 lethality might be higher in younger cases. Special attention should be given to susceptible individuals and screening should be conducted for all symptomatic cases with either obesity and/or diabetes to decrease the burden associated with COVID-19 in Mexico.

## Data Availability

All data sources and R code are available for reproducibility of results at https://github.com/oyaxbell/covid_diabetesmx

## Conflict of interest

Nothing to disclose.

## Role of the funding source

This research received no funding.

## ACKNOWLEDGMENTS

JPBL, AVV, NEAV, AMS and CAFM are enrolled at the PECEM program of the Faculty of Medicine at UNAM. JPBL and AVV are supported by CONACyT. The authors would like to acknowledge the invaluable work of all of Mexico’s healthcare community in managing the COVID-19 epidemic. Its participation in the COVID-19 surveillance program has made this work a reality, we are thankful for your effort.

## DATA AVAILABILITY

All data sources and R code are available for reproducibility of results at https://github.com/oyaxbell/coviddiabetesmx.

## AUTHOR CONTRIBUTIONS

Research idea and study design OYBC, JPBL, CAAS; data acquisition: OYBC, AGD; data analysis/interpretation: OYBC, JPBL, NEAV, AVV, AGD, JJN, CAAS; statistical analysis: OYBC, NEAV; manuscript drafting: OYBC, NEAV, AVV, JPBL, AMS, CAFM, JJN; supervision or mentorship: OYBC, CAAS. Each author contributed important intellectual content during manuscript drafting or revision and accepts accountability for the overall work by ensuring that questions pertaining to the accuracy or integrity of any portion of the work are appropriately investigated and resolved.

## FUNDING

No funding was received.

## CONFLICT OF INTEREST/FINANCIAL DISCLOSURE

Nothing to disclose.

## Notes

**CONFLICT OF INTERESTS:** Nothing to disclose.

### Competing Interest Statement

The authors have declared no competing interest.

### Funding Statement

No funding received.

### Author Declarations

No approval necessary.

